# Association between maternal genome-wide polygenic scores for psychiatric and neurodevelopmental disorders and perinatal risk factors: A Danish population-based study

**DOI:** 10.1101/2025.01.15.25320615

**Authors:** Fenfen Ge, Yue Wang, Xiaoqin Liu, Trine Munk-Olsen, Kathrine Bang Madsen, Emil Michael Pedersen, Clara Albiñana, Esben Agerbo, Cynthia M. Bulik, Liselotte Vogdrup Petersen, Unnur A. Valdimarsdottir, Bjarni Jóhann Vilhjálmsson

**Affiliations:** Centre of Public Health Sciences, Faculty of Medicine, University of Iceland, Reykjavík, Iceland; NCRR-The National Centre for Register-based Research, Department of Public Health, Aarhus University, Aarhus, Denmark; Child and Adolescent Psychiatry Research Unit, Department of Clinical Research, University of Southern Denmark, Odense, Denmark; Department of Medical Epidemiology & Biostatistics, Karolinska Institutet, Stockholm, Sweden; Department of Psychiatry, School of Medicine, University of North Carolina at Chapel Hill, Chapel Hill, USA; Department of Nutrition, University of North Carolina at Chapel Hill, Chapel Hill, USA; Harvard T.H. Chan School of Public Health, Boston, United States; Bioinformatics Research Centre, Aarhus University, Aarhus, Denmark; Novo Nordisk Foundation Centre for Genomic Mechanisms of Disease, the Broad Institute of MIT and Harvard, Cambridge, MA, USA

## Abstract

**Importance:** Adverse perinatal outcomes are common challenges for mothers and their newborns. Epidemiological studies indicate that mothers with psychiatric and neurodevelopmental disorders are at an increased risk of adverse pregnancy and neonatal outcomes; however, the underlying mechanisms behind these associations remain inadequately understood.

**Objective:** To investigate whether perinatal risk factors are driven by maternal genetic susceptibility to multiple psychiatric and neurodevelopmental disorders.

**Design, setting and participants:** This nationwide population-based case-control study identified 14,917 primiparous mothers with available genetic information, born between 1981 and 2008, from the iPSYCH cohort, which is nested in the Danish National Registers.

**Exposures:** Eight genome-wide polygenic scores (PGS) for psychiatric and neurodevelopmental disorders in mothers—ADHD, autism spectrum disorder (ASD), schizophrenia, depression, anxiety, bipolar disorder, obsessive-compulsive disorder, and anorexia nervosa—were calculated using LDPred2.

**Main outcomes and measures:** Six pregnancy-related and four neonatal-related risk factors were obtained from the Danish Medical Birth Registry. Odds ratios (ORs) and 95% CIs were estimated, adjusted for the year of delivery and the first 10 genetic principal components.

**Results:** Of 14,917 mothers, the mean age at childbirth was 24.9 years (standard deviation [SD]=3.9). Per SD increase in the PGS for ADHD (OR=1.07 [95%CI 1.00-1.14]), anxiety (1.10 [1.03-1.18]) and depression (1.12 [1.05-1.20]) were associated with maternal smoking exceeding 10 cigarettes per day during the early pregnancy. Stronger associations were observed for the depression PGS in relation to younger age at first birth (i.e. < 20 years; 1.15 [1.07-1.23]), mandatory education (1.15 [1.11-1.19]), and non-cohabitation status during pregnancy (1.07 [1.02-1.12]), compared to other PGS. Schizophrenia PGS was associated with reduced odds of maternal obesity (0.88 [0.84-0.93]) during early pregnancy. In contrast, little evidence was found for associations between maternal PGS for psychiatric and neurodevelopmental disorders and neonatal-related risk factors. We observed comparable associations when the analyses excluded mothers with any psychiatric or neurodevelopmental disorders prior to the conception date.

**Conclusions and relevance:** High genetic loading for psychiatric and neurodevelopmental disorders may partly explain the observed phenotypic associations between maternal mental illness and perinatal risk factors, particularly pregnancy related factors, but it is less likely to account for associations with neonatal related factors. Alternative mechanisms, e.g., psychological stress and medical treatment for psychiatric and neurodevelopmental disorders, should be further explored.

## Introduction

Perinatal risk factors, such as preterm birth, small for gestational age, and younger or advanced maternal age at childbirth are among the leading causes of short- and long-term health issues for both mother and newborn.^1–3^ Accumulating epidemiological evidence based on large-scale prospective national health registers or nationwide cohorts has shown that women with pre-existing or active psychiatric and neurodevelopmental disorders (e.g., schizophrenia,^4^ anorexia nervosa,^5^ obsessive-compulsive disorder,^6^ bipolar disorder,^7^ depression,^7,8^ or autism spectrum disorder^9^) are at increased risk of a variety of adverse pregnancy behaviours (e.g., more frequent smoking during pregnancy,^4^ and caesarean sections^10^) and neonatal outcomes (e.g., preterm birth,^5–7^ low Apgar at 5 minutes,^6,10^ and low birthweight^7^). A recent meta-analysis on 43,611 deliveries of women with schizophrenia and 40,948,272 controls across 11 high-income countries found that schizophrenia was associated with a substantially increased risk of very preterm delivery and stillbirth.^11^ However, the underlying mechanisms driving these associations remain inadequately understood, with unmeasured confounding by shared genetic or environmental factors potentially contributing to the observed associations.

Genetic factors play an important role in the development of mental illness, as shown in genome-wide associations studies (GWAS) and twin studies.^12,13^ Specifically, estimated SNP-based heritability on the liability scale is 8.9% for depression,^14^ 10% for anxiety,^15^ 11-17% for anorexia nervosa,^16^ and 24% for schizophrenia^17^ with hundreds or thousands of genetic loci identified via GWAS. Polygenic scores (PGS), which combine the effect sizes of multiple genetic variants using well-established summary data, can be used to assess genetic liability of an individual to specific mental illnesses, such as schizophrenia or depression.^18^ Understanding the role of genetic factors for maternal mental illness in perinatal risk factors is important for improved understanding of factors influencing adverse pregnancy and neonatal outcomes.

Yet, evidence on the associations between maternal genetic liability to mental illnesses and perinatal risk factors is lacking, likely due to the scarcity of maternal genetic information or well-documented pregnancy and birth characteristics.^19–21^ Moreover, previous studies have not had sufficient long follow-up periods to assess the pre-pregnancy mental health history of the mothers, limiting the ability to exclude potential phenotypic effects of mental illness that has already manifested and to disentangle those factors from the underlying impact of maternal genetic liability to psychiatric and neurodevelopmental disorders. To this end, leveraging individual genotyping data from the Integrative Psychiatric Research (iPSYCH) cohort and detailed pregnancy, delivery, and birth information from the Danish National Registers, we aimed to assess the associations between eight genome-wide polygenic scores for psychiatric and neurodevelopmental disorders for mother and perinatal risk factors (six pregnancy-related factors and four neonatal-related factors).

## Methods

### Date Source and study population

We used data from the Integrative Psychiatric Research (iPSYCH2015) study,^22^ a population-based case-cohort sample that builds upon the design of iPSYCH2012.^23^ The iPSYCH2015 sample was selected from the Civil Registration System, which includes all singletons born between 1981 and 2008 who were alive and living in Denmark at one year of age, with known maternal information. A total of 93,608 individuals diagnosed with one or more major psychiatric disorders were identified through the Danish Psychiatric Central Research Register and included in the case group. The anorexia nervosa (AN; ANGI-DK) samples were include from the Anorexia Nervosa Genetics Initiative (ANGI),^24^ as they were samples within the same framework as iPSYCH 2015. Moreover, a random sample of 50,615 individuals from the same birth cohort was selected to serve as a population-representative control group. DNA was extracted from blood samples collected at birth and stored as dried blood spots in the Danish Newborn Screening Biobank.^25^ Detailed information about iPSYCH2015 has been described in the cohort profile.^22^

Information on demographic characteristics (e.g., birth year or sex), medical diagnoses (e.g., ICD-10 code), prescription and birth information were derived by cross-linkage with Danish national registers including, Medical Birth register,^26^ Psychiatric Central Research Register,^27^ National Prescription Registry,^28^ and Civil Registration System.^29,30^ Detailed descriptions of registers used are shown in Supplementary table 1.

In the present study, among all individuals in the iPSYCH2015, we first excluded individuals who did not pass genotypic quality control,^31^ showed significant heterogeneity (≥4.5 log distance units), or had a degree of relatedness greater than second-degree (0.0884), leaving 108,628 for further analysis. We then linked data from iPSYCH2015 to the Medical Birth register and included 14,917 primiparous mothers giving birth to singletons in the analysis. To examine the direct pathway of maternal psychiatric genetic susceptibility, we further excluded mothers with any psychiatric disorder diagnosis (ICD-10 F codes) or records of antidepressant use (ATC code N06A) from mother’s birth through six months prior to conception and included 7,816 mothers for analysis (**Figure 1**).

**Figure 1.**
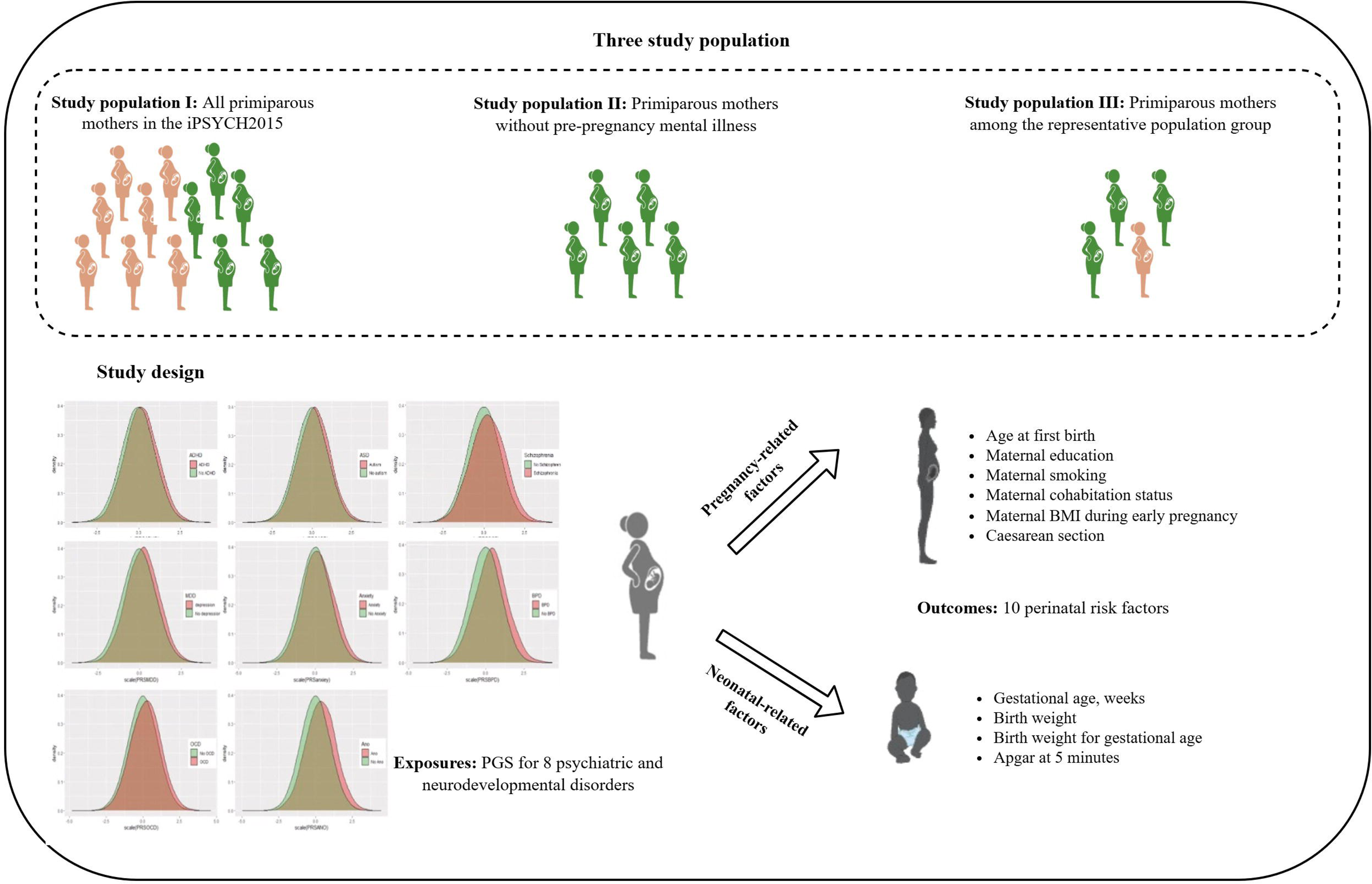
Study profile

The present study obtained approval from the Danish Scientific Ethics Committee, the Danish Health Data Authority, the Danish Data Protection Agency, and the Danish Neonatal Screening Biobank Steering Committee.

### Genotyping

Standard GWAS quality control procedures were applied in iPSYCH2015, which involved removing SNPs with a minor allele frequency (MAF) < 0.01 and a Hardy-Weinberg equilibrium p-value < 10⁻⁶, as well as restricting the analysis to HapMap3 variants in the LD reference panel.^31^ This resulted in a final dataset of 1,053,299 SNPs. Principal component analysis (PCA) was conducted, yielding 20 principal components (PCs) and genetically homogeneous individuals were defined as those with a log distance of <4.5 units from the multidimensional centre of the 20 PCs.^32^ The KING-relatedness robust coefficient was estimated and used to exclude participants with a degree of relatedness greater than second-degree (kinship > 0.0884).^33^

### Polygenic score for psychiatric and neurodevelopmental disorders

We generated polygenic scores (PGS) for ADHD, ASD, schizophrenia, depression, anxiety, bipolar disorder, obsessive-compulsive disorder (OCD), and anorexia nervosa using the GWAS summary statistics from the Psychiatric Genomics Consortium, excluding iPSYCH2015 sample, with the LDpred2 method.^34^ Detailed information about summary statistics and validation information for each PGS was shown in our previous study and Supplementary Table 2.^35^ The PGS were standardised to the mean and standard deviation of the study population in the analysis. The distribution of the standardized PGSs of interest are shown in Supplementary Figure 1. The PGS for mothers without pre-pregnancy diagnosed psychiatric or neurodevelopmental disorders or antidepressant use was slightly lower than that of mothers with such diagnoses or medication use.

### Measurements of perinatal risk factors

We categorised factors during pregnancy and delivery, pertaining to the mother, as pregnancy-related risk factors, and those concerning the newborn and the time surrounding delivery as neonatal-related risk factors.

#### Pregnancy-related factors

Information on maternal smoking (categorised as non-smoker, stopping smoking, 1-10 cigarettes/day, and greater than 10 cigarettes/day) and maternal BMI during early pregnancy (categorised as <25 [underweight and normal], 25 to 30 [overweight], and ⩾30 kg/m² [obesity]) was derived from the Medical Birth Register. Age at first birth was calculated by subtracting birth date of first baby derived from Medical Birth Register from birth date of the mother derived from civil registration system, and categorised as <20, 20-24, 25-29, and 30-36 years. Information on both maternal cohabitation status (i.e., living with a partner or not) and maternal education (i.e., mandatory or above mandatory) during pregnancy was extracted from the Medical Birth Register and the civil registration system, respectively. The caesarean section was recorded in Medical Birth Register and grouped as ‘with’ or ‘without’.

#### Neonatal-related factors

Gestational age was estimated from the ultrasound scan; if unknown, 280 days were used as a replacement.^36^ It was then categorised as preterm (≤36 weeks), term (36-41 weeks), or post-term (>41 weeks).^37^ We first classified birth weight as <2500g, 2500-3999g, and >3999g. We then calculated birth weight for gestational age using sex-specific reference curves for foetal growth^38^ and categorized into <10^th^ percentile (small for gestational age), 10^th^ to 90^th^ percentile (normal for gestational age), and >90^th^ percentile (large for gestational age). The Apgar score at five minutes was used and categorized into two groups: 7-10 for normal and <7 for low.^39^

### Covariates

Information on the calendar year of delivery was obtained from the Medical Birth Register and treated as a continuous variable, ranging from 1981 to 2008. Additionally, the first 10 standardized genetic principal components were adjusted to account for population substructure in the genetic data.^23^

### Statistical analysis

#### Main analyses

Descriptive results are reported as frequencies and percentages. We assessed the associations between eight types of PGS and perinatal risk factors, using logistic regression models for binary variables (e.g., maternal cohabitation status) or multinomial logistic regression models for categorical variables (e.g., maternal age at first birth). The results were represented as odds ratios (ORs) and 95% confidence intervals (CIs), and the effect estimates are presented per 1-standard deviation (1-SD) increase in PGS to aid interpretation. The analyses included the calendar year of delivery to account for genotyping waves and the first 10 standardized genetic principal components to adjust for population substructure. All data analyses were conducted using R-4.0 software and the statistical significance was adjusted for multiple testing using the Bonferroni correction.^40^

#### Secondary analyses

Since the population controls were randomly selected from the entire Danish population,^23^ we re-ran the analyses restricted to primiparous mothers within the representative population group to obtain estimates more reflective of the national population (**Figure 1**). We performed a negative control analysis using left-handedness PGS, as left-handedness has not been associated with any perinatal risk factors.^41^

## Results

Of 14,917 mothers included in the present study, the mean age at first birth was 24.9 years (standard deviation, 3.9 years). Compared to the population representative group, the entire included population was more likely to have a younger maternal age at first birth (i.e., <20 years), standard education level (i.e., mandatory education), and higher rates of non-cohabitation and smoking during the early pregnancy (**Table 1**).

**Table 1.**
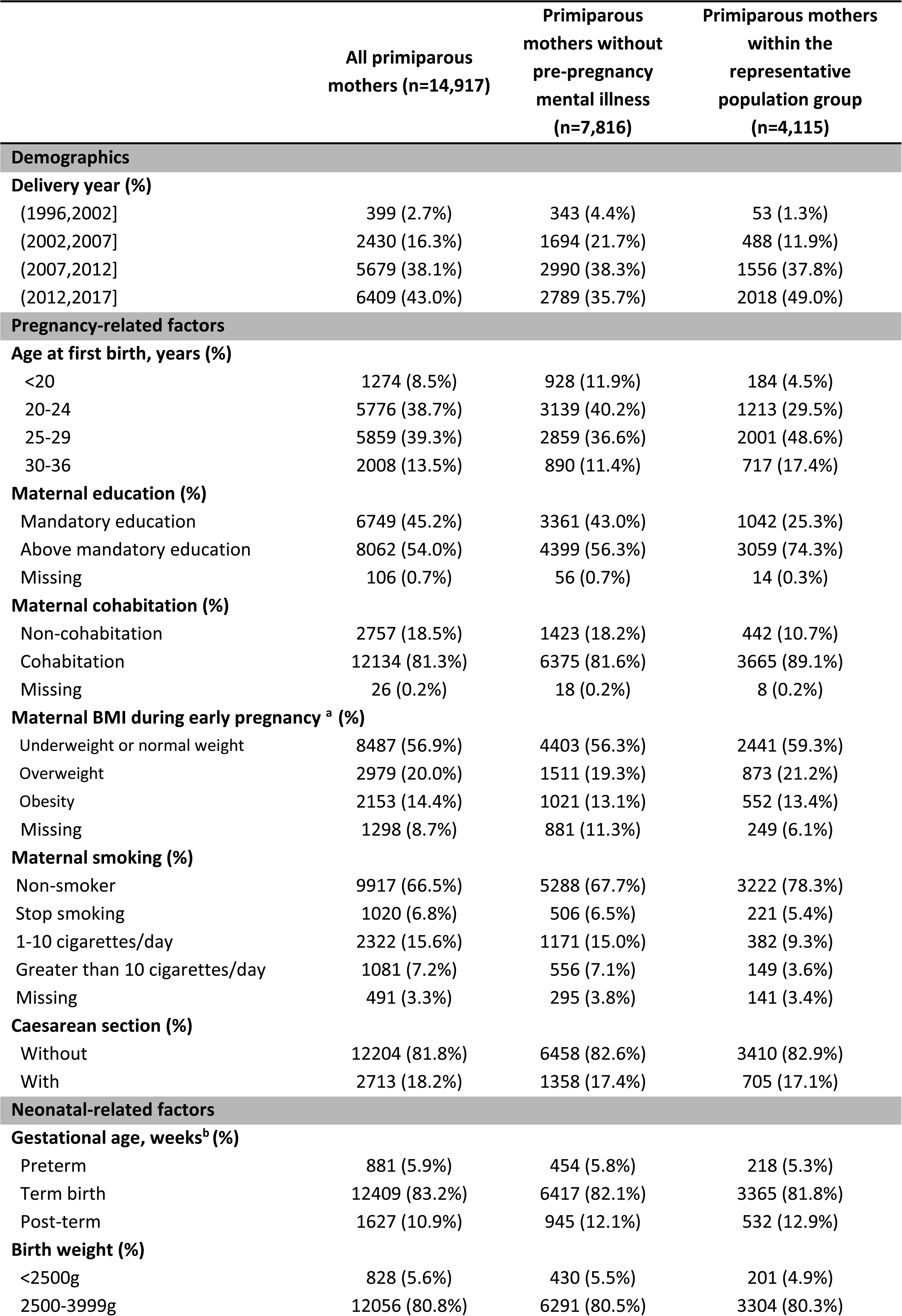
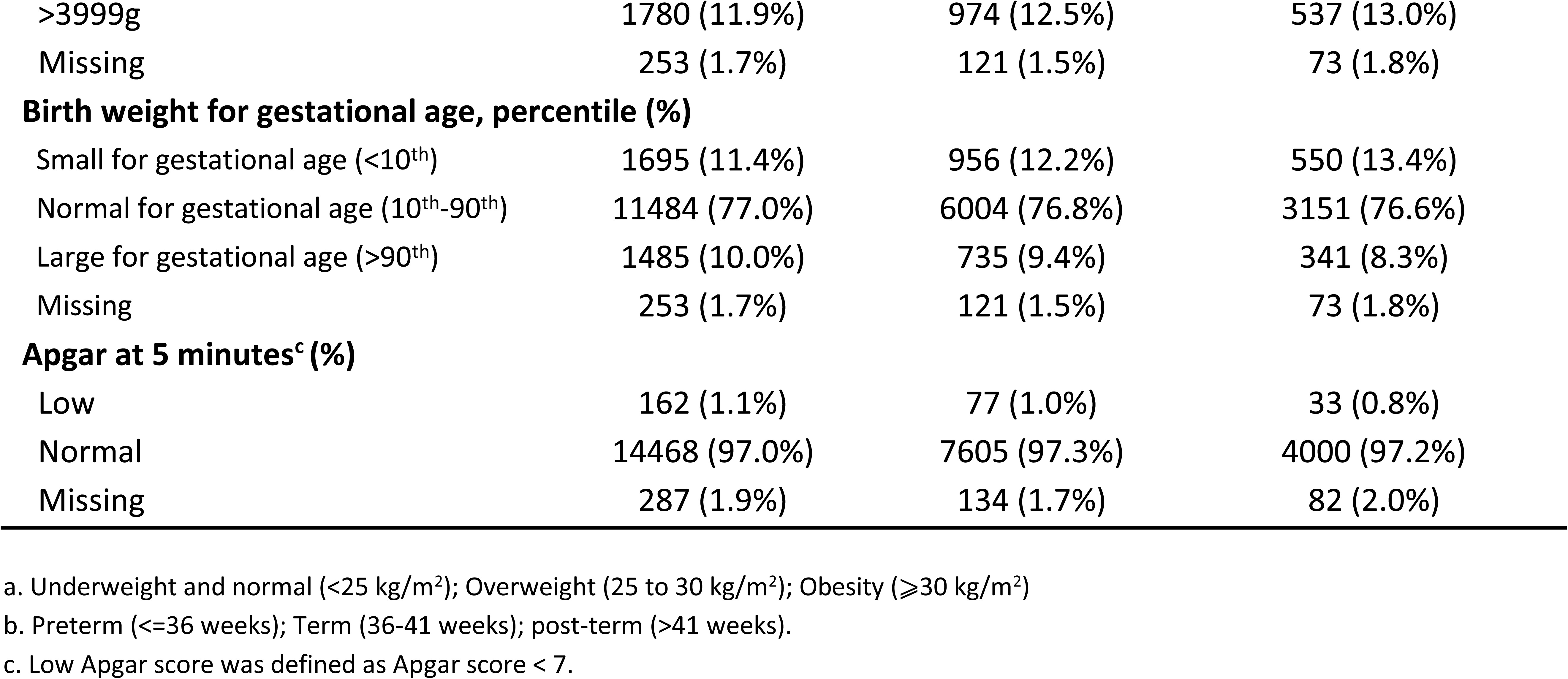
Characteristics of study population.

### Pregnancy-related risk factors

Associations between each maternal psychiatric or neurodevelopmental PGS and pregnancy-related risk factors are shown in **Figure 2**. PGS for ADHD (OR=1.07 [95%CI 1.00-1.14]), anxiety (1.10 [1.03-1.18]), and depression (1.12 [1.05-1.20]) were associated with maternal smoking during pregnancy exceeding 10 cigarettes/day. PGS for ADHD (1.05 [1.02-1.09]) and anxiety (1.07 [1.03-1.11]) were associated with standard education level during pregnancy, and PGS for schizophrenia (1.08 [1.03-1.12]) was associated with non-cohabitation status during pregnancy.

**Figure 2.**
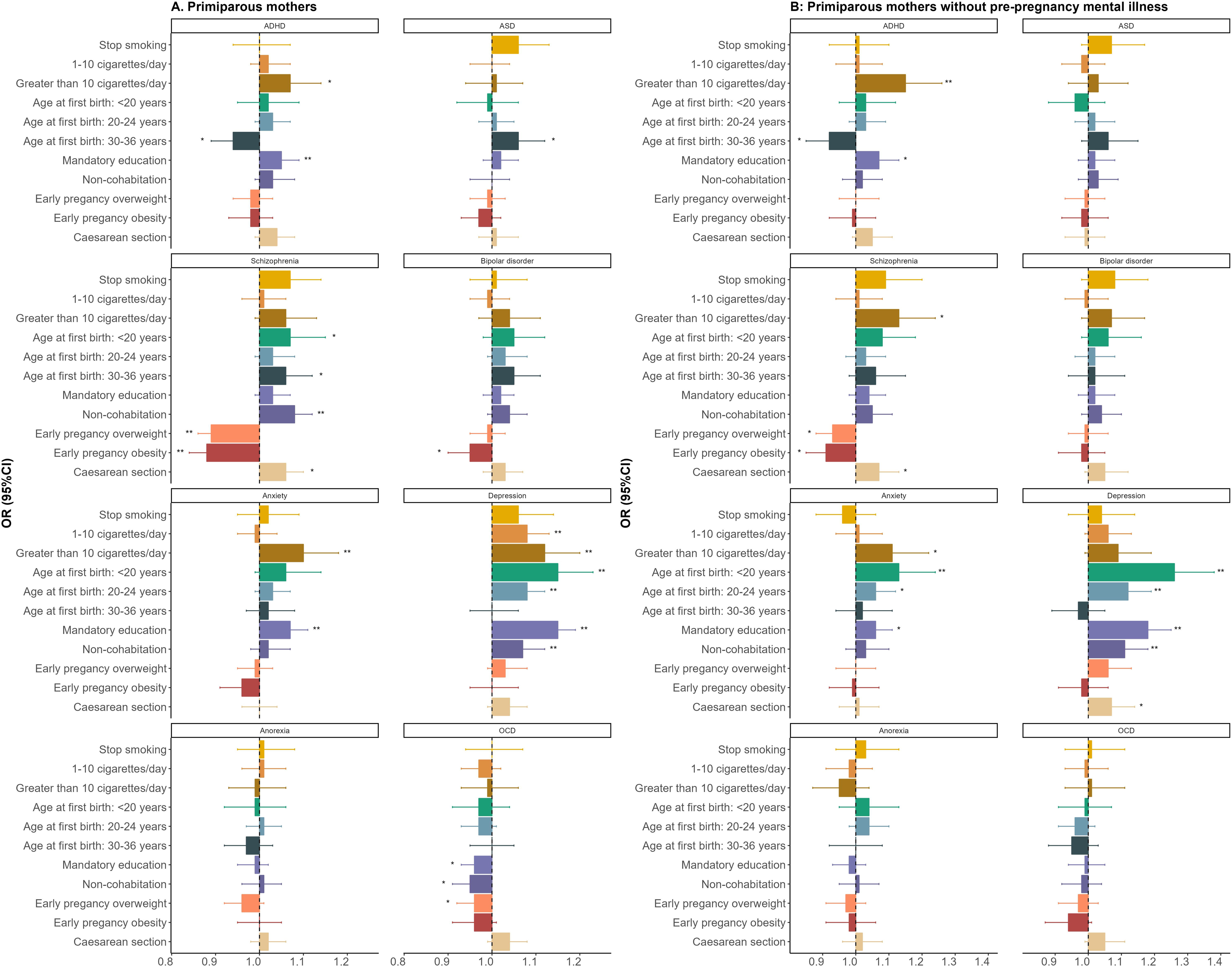
Associations between maternal polygenic scores for psychiatric and neurodevelopmental disorders and pregnancy-related factors. * p < 0.05; ** p.adjust < 0.0083

In contrast, stronger associations with younger age at first birth (i.e., < 20 years; 1.15 [1.07-1.23]), standard education level (1.15 [1.11-1.19]), and non-cohabitation status during pregnancy (1.07 [1.02-1.12]) were found for the depression PGS, and the observed association for depression PGS remained significant when restricted to mothers without diagnosed pre-pregnancy mental illness.

The schizophrenia PGS was associated with reduced odds of being having maternal overweight (0.89 [0.86-0.93]) and obesity (0.88 [0.84-0.93]) during early pregnancy. Similar results were observed among mothers without diagnosed pre-pregnancy mental illness, although these were not significant after correcting for multiple testing.

No or weak associations were found between the PGS for ASD, bipolar disorder, OCD, and anorexia nervosa and pregnancy-related risk factors, both in the full sample of included primiparous mothers and in mothers without diagnosed pre-pregnancy mental illness.

### Neonatal-related risk factors

Associations between each maternal psychiatric or neurodevelopmental PGS and neonatal-related outcomes are shown in **Figure 3**. Except for an observed association between PGS for anxiety and preterm birth (0.91 [0.84-0.97]), low birth weight (<2500g; 0.91 [0.84-0.97]), and being small for gestational age (<10th percentile, 0.93 [0.89-0.98]), no associations were found between other maternal psychiatric or neurodevelopmental PGS and neonatal-related factors. No results survive multiple testing when restricted to mothers without pre-pregnancy mental illness.

**Figure 3.**
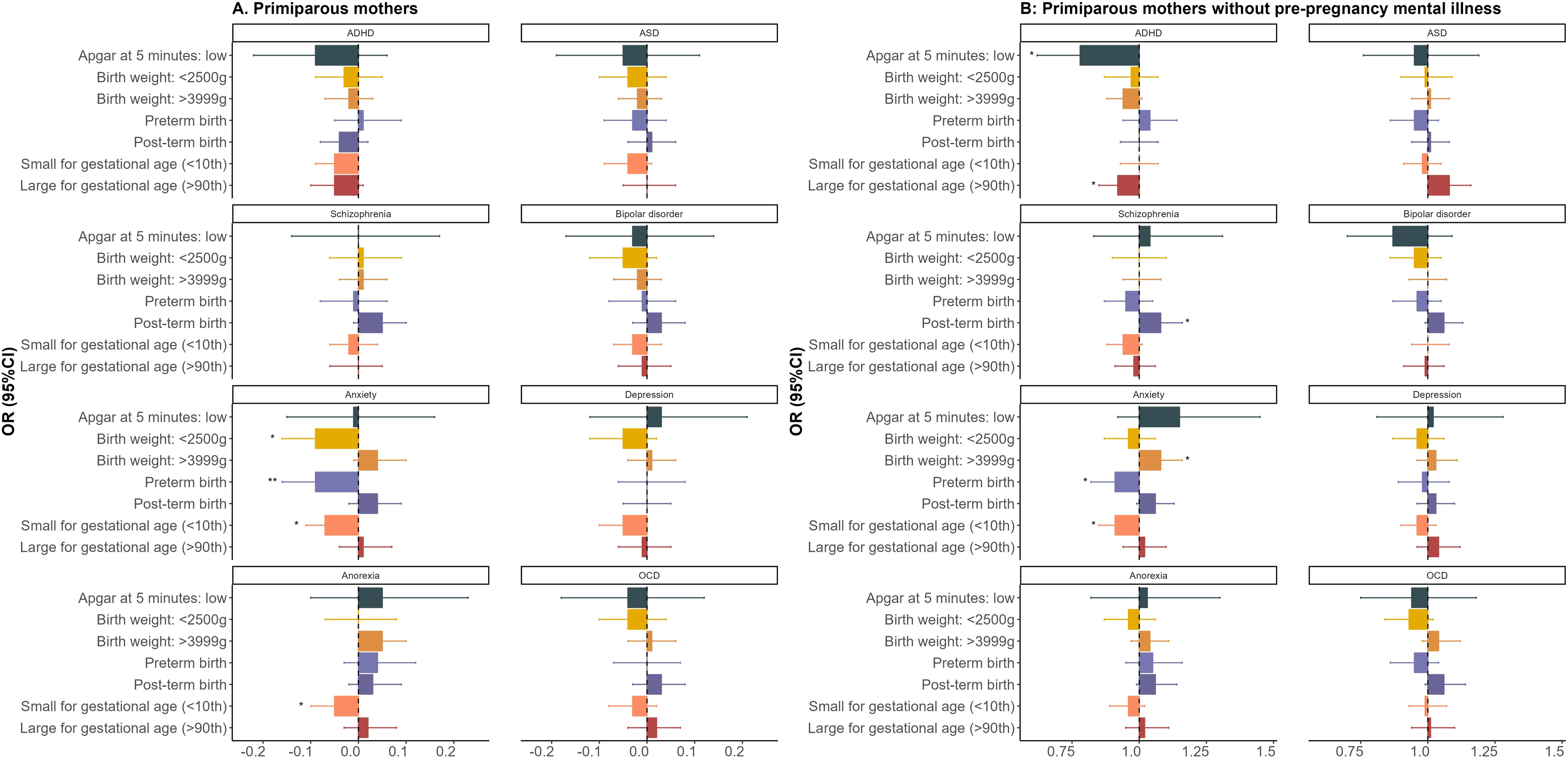
Associations between maternal polygenic scores for psychiatric and neurodevelopmental disorders and neonatal-related factors. * p < 0.05; ** p.adjust < 0.0125

### Secondary analyses

Largely comparable results were observed among the representative population group (Supplementary Figure 2). As expected, no statistically significant associations were observed between negative control exposure (PGS for left-handedness) and any pregnancy- or neonatal-related risk factors (Supplementary Table 3).

## Discussion

This study examined the association between maternal genetic liability—measured by eight psychiatric and neurodevelopmental disorder-specific PGS—and a range of perinatal risk factors. The results of our study indicate a link between maternal psychiatric or neurodevelopmental PGS and pregnancy related risk factors, including maternal smoking during pregnancy, younger age at first birth, non-cohabitation during pregnancy, standard education level, and a reduced likelihood of being overweight in early pregnancy. Moreover, we found that part of this association, particularly for depression PGS, persisted even when the analysis was restricted to mothers without diagnosed pre-pregnancy mental illness. In contrast, our findings do not support a link between maternal psychiatric or neurodevelopmental PGS and neonatal-related risk factors.

Our maternal smoking results aligned with previous findings from the Norwegian Mother, Father, and Child Cohort Study (MoBa),^20^ the Avon Longitudinal Study of Parents and Children (ALSPAC),^42^ and the Nurses’ Health Study 2 (NHS2).^19^ We extended prior epidemiological studies^43,44^ and found that mothers with a high genetic risk for the ADHD, anxiety, and depression were more likely to smoke heavily during pregnancy. However, no association was noted between schizophrenia PGS and maternal smoking, which was inconsistent with findings from a previous study, suggesting that this discrepancy may be attributed to differences in the definition of the outcome.^19^ Our results further suggest that maternal PGS, particularly for depression, is associated with a younger age at first birth, standard education level, and non-cohabitation during pregnancy. Expanding upon previous studies using both linkage disequilibrium score regression (LDSC)^45^ and Mendelian randomization (MR) analyses,^46^ we found that the associations persisted even among mothers with no diagnosed pre-pregnancy mental illness, suggesting a direct effect of maternal genetic liability to psychiatric disorders. For example, one study suggested associations of age at first birth and depression using univariable MR,^47^ although another study did not totally support these findings based on multivariable MR when considering age at first sexual intercourse, first birth, last birth, and age at menopause.^48^ Compared to methods that utilise summary data from GWAS (e.g., LDSC or MR) to calculate genetic correlations, associations derived from PGS offer specific indices relevant to aetiology, potentially improving the utility of PGS testing in clinical settings.^49^

Both the negative associations between schizophrenia PGS and maternal overweight/obesity in early pregnancy and the absence of associations between other types of genetic risk for psychiatric and neurodevelopmental disorders (i.e. ADHD, ASD, anxiety, depression, bipolar disorder, anorexia nervosa, and OCD) and overweight/obesity in our study are novel. Indeed, mounting evidence suggest higher rates of overweight/obesity in individuals with mental illnesses, such as schizophrenia, bipolar disorder, and depression.^50^ However, the findings of our study, and the Zhang et al. study,^49^ based on UK Biobank data of 406, 929 individuals without a schizophrenia diagnosis, do not support the positive genetic associations between schizophrenia PGS and overweight or obesity. Psychiatric disorders are commonly recognized as chronic or recurrent conditions, often requiring maintenance therapy involving mood stabilisers, anxiolytics, antidepressants, or antipsychotics, all of which can cause weight gain by influencing appetite control or energy metabolism.^51^ Thus, the effects of psychiatric medication might partly explain the aforementioned phenotypic association.

Consistent with previous findings from ALSPAC^42^ and NHS2^19^, we found little evidence of an association between maternal PGS for psychiatric or neurodevelopmental disorders and neonatal related risk factors, including birth weight and gestational age. Similarly, the study using data from the ALSPAC^42^ suggested that neither maternal nor child neurodevelopmental genetic liability (i.e., ADHD, and ASD) were associated with low birth weight, preterm delivery, or low Apgar score at 5 minutes. Instead of maternal genetic liability to these disorders, factors such as psychotropic medication and psychological stress may explain the discrepancy between observed phenotypic associations and absent genetic associations. For example, several previous meta-analyses have found that the use of antidepressants during pregnancy is associated with an increased risk of preterm birth and low birth weight, regardless of whether the comparison group consists of all unexposed mothers or only depressed mothers without antidepressants.^52–54^ Moreover, a meta-analysis that included 23 studies found that women with depression who were not receiving treatment for their depression had significantly increased infant risks of preterm birth and low birth weight compared to pregnant women without depression.^55^ Thus, these findings suggest that medication may not be the only contributing factor, psychological stress, such as guilt or stigma associated with mental illness,^56^ may mediate these associations.

### Strengths and Limitations

To the best of our knowledge, this is the first study to comprehensively explore the associations between eight types of psychiatric and neurodevelopmental PGS and well-documented perinatal risk factors among primiparous mothers. Another major strength of our study is the restriction of analyses to mothers without any pre-existing mental illnesses, which allows for direct exploration of whether associations between maternal mental illnesses and pregnancy and neonatal-related risk factors are driven by genetic liability. There are several limitations that need to be considered. First, the iPSYCH2015 cohort includes individuals born between 1981 and 2008, with the Medical Birth Register updated until 2017. As a result, our study primarily captures relatively young mothers (average: aged 24.9 years), which may lead to systematic differences in the associations between genetic risk for mental illness and the outcomes tested. Some mother may not yet have traversed the age of risk for developing some of the disorders studied. Second, despite the large sample size, our study lacks sufficient power to investigate pregnancy-related conditions such as gestational hypertension and diabetes, given the predominance of younger mothers in the cohort. Thirdly, maternal smoking during the pregnancy was self-reported in our study, and we used a detailed classification based on the number of cigarettes smoked per day. While this provides more detailed information, it may introduce information bias compared to a simpler ‘Yes’ or ‘No’ classification. Additionally, it should be noted that, in our study, the maternal BMI in early pregnancy was assessed at early stage of pregnancy and nearly 11% of the data is missing. Finally, while our study identifies associations between PGS and pregnancy-related risk factors, it does not infer causal relationships between maternal genetic liability and these factors. Future research is needed to determine the causal direction of the relationship between them.

## Conclusions

Within a large cohort covering a total of 14,917 primiparous mothers, we found that genetic risk may partly account for previously identified associations between maternal psychiatric and neurodevelopmental disorders and pregnancy-related risk factors, even in the absence of diagnosed disorders. In contrast, we found the associations between maternal mental illness and neonatal-related risk factors are less likely driven by the maternal genetic liability to psychiatric or neurodevelopmental disorders. Future studies could replicate our findings in larger, well-powered datasets and incorporate explorations of additional mechanisms, such as psychological stress or psychopharmacological treatment on maternal and neonatal outcomes.

## Data Availability

Data availability is limited due to the sensitive nature. For more information please contact the authors.

## Acknowledgements

The authors gratefully acknowledge the Psychiatric Genomics Consortium (PGC) and the research participants and employees of 23andMe, Inc., for providing the summary statistics used to generate the polygenic score.

## Reference

1. Vandekerckhove M, Guignard M, Civadier M-S, Benachi A, Bouyer J. Impact of maternal age on obstetric and neonatal morbidity: a retrospective cohort study. BMC pregnancy and childbirth. 2021;21:1–7.

2. Crump C, Sundquist J, Sundquist K. Preterm delivery and long term mortality in women: national cohort and co-sibling study. BMJ. 2020;370:m2533. doi:10.1136/bmj.m2533

3. Yang F, Janszky I, Gissler M, et al. Preterm Birth, Small for Gestational Age, and Large for Gestational Age and the Risk of Atrial Fibrillation Up to Middle Age. JAMA Pediatrics. 2023;177(6):599–607. doi:10.1001/jamapediatrics.2023.0083

4. Fabre C, Pauly V, Baumstarck K, et al. Pregnancy, delivery and neonatal complications in women with schizophrenia: a national population-based cohort study. Lancet Reg Health Eur. Nov 2021;10:100209. doi:10.1016/j.lanepe.2021.100209

5. Mantel Ä, Hirschberg AL, Stephansson O. Association of Maternal Eating Disorders With Pregnancy and Neonatal Outcomes. JAMA Psychiatry. Mar 1 2020;77(3):285–293.

6. Fernandez de la Cruz L, Joseph KS, Wen Q, Stephansson O, Mataix-Cols D, Razaz N. Pregnancy, Delivery, and Neonatal Outcomes Associated With Maternal Obsessive-Compulsive Disorder: Two Cohort Studies in Sweden and British Columbia, Canada. JAMA network open. Jun 1 2023;6(6):e2318212. doi:10.1001/jamanetworkopen.2023.18212

7. Lee HC, Lin HC. Maternal bipolar disorder increased low birthweight and preterm births: a nationwide population-based study. J Affect Disord. Feb 2010;121(1-2):100–5. doi:10.1016/j.jad.2009.05.019

8. De Mola CL, De França GVA, de Avila Quevedo L, Horta BL. Low birth weight, preterm birth and small for gestational age association with adult depression: systematic review and meta-analysis. British Journal of Psychiatry. 2014;205(5):340–347. doi:10.1192/bjp.bp.113.139014

9. Hosozawa M, Cable N, Ikehara S, et al. Maternal Autistic Traits and Adverse Birth Outcomes. JAMA network open. Jan 2 2024;7(1):e2352809.

10. Momen NC, Chatwin H, Holde K, et al. Maternal mental disorders and neonatal outcomes: Danish population-based cohort study. Br J Psychiatry. Oct 8 2024:1–8. doi:10.1192/bjp.2024.164

11. Etchecopar-Etchart D, Mignon R, Boyer L, Fond G. Schizophrenia pregnancies should be given greater health priority in the global health agenda: results from a large-scale meta-analysis of 43,611 deliveries of women with schizophrenia and 40,948,272 controls. Mol Psychiatry. Aug 2022;27(8):3294–3305. doi:10.1038/s41380-022-01593-9

12. Pettersson E, Lichtenstein P, Larsson H, et al. Genetic influences on eight psychiatric disorders based on family data of 4 408 646 full and half-siblings, and genetic data of 333 748 cases and controls. Psychol Med. May 2019;49(7):1166–1173.

13. Collins AL, Sullivan PF. Genome-wide association studies in psychiatry: what have we learned? The British journal of psychiatry: the journal of mental science. Jan 2013;202(1):1–4.

14. Howard DM, Adams MJ, Clarke TK, et al. Genome-wide meta-analysis of depression identifies 102 independent variants and highlights the importance of the prefrontal brain regions. Nature neuroscience. Mar 2019;22(3):343–352. doi:10.1038/s41593-018-0326-7

15. Otowa T, Hek K, Lee M, et al. Meta-analysis of genome-wide association studies of anxiety disorders. Mol Psychiatry. Oct 2016;21(10):1391–9. doi:10.1038/mp.2015.197

16. Watson HJ, Yilmaz Z, Thornton LM, et al. Genome-wide association study identifies eight risk loci and implicates metabo-psychiatric origins for anorexia nervosa. Nat Genet. Aug 2019;51(8):1207–1214.

17. Trubetskoy V, Pardinas AF, Qi T, et al. Mapping genomic loci implicates genes and synaptic biology in schizophrenia. Nature. Apr 2022;604(7906):502-508. doi:10.1038/s41586-022-04434-5

18. Lewis CM, Vassos E. Polygenic Scores in Psychiatry: On the Road From Discovery to Implementation. Am J Psychiatry. Nov 1 2022;179(11):800–806. doi:10.1176/appi.ajp.20220795

19. Ratanatharathorn A, Chibnik LB, Koenen KC, Weisskopf MG, Roberts AL. Association of maternal polygenic risk scores for mental illness with perinatal risk factors for offspring mental illness. Sci Adv. Dec 14 2022;8(50):eabn3740.

20. Havdahl A, Wootton RE, Leppert B, et al. Associations Between Pregnancy-Related Predisposing Factors for Offspring Neurodevelopmental Conditions and Parental Genetic Liability to Attention-Deficit/Hyperactivity Disorder, Autism, and Schizophrenia: The Norwegian Mother, Father and Child Cohort Study (MoBa). JAMA Psychiatry. Aug 1 2022;79(8):799–810. doi:10.1001/jamapsychiatry.2022.1728

21. Wiegersma AM, Dalman C, Lee BK, Karlsson H, Gardner RM. Association of Prenatal Maternal Anemia With Neurodevelopmental Disorders. JAMA Psychiatry. Dec 1 2019;76(12):1294–1304. doi:10.1001/jamapsychiatry.2022.1728

22. Bybjerg-Grauholm J, Bøcker Pedersen C, Bækvad-Hansen M, et al. The iPSYCH2015 Case-Cohort sample: updated directions for unravelling genetic and environmental architectures of severe mental disorders. medRxiv. 2020:2020.11. 30.20237768.

23. Pedersen CB, Bybjerg-Grauholm J, Pedersen MG, et al. The iPSYCH2012 case-cohort sample: new directions for unravelling genetic and environmental architectures of severe mental disorders. Mol Psychiatry. Jan 2018;23(1):6–14. doi:10.1038/mp.2017.196

24. Thornton LM, Munn-Chernoff MA, Baker JH, et al. The Anorexia Nervosa Genetics Initiative (ANGI): Overview and methods. Contemp Clin Trials. Nov 2018;74:61–69. doi:10.1016/j.cct.2018.09.015

25. Nørgaard-Pedersen B, Hougaard DM. Storage policies and use of the Danish Newborn Screening Biobank. J Inherit Metab Dis. Aug 2007;30(4):530–6. doi:10.1007/s10545-007-0631-x

26. Bliddal M, Broe A, Pottegård A, Olsen J, Langhoff-Roos J. The Danish Medical Birth Register. European Journal of Epidemiology. 2018/01/01 2018;33(1):27–36. doi:10.1007/s10654-018-0356-1

27. Mors O, Perto GP, Mortensen PB. The Danish Psychiatric Central Research Register. Scand J Public Health. Jul 2011;39(7 Suppl):54-7. doi:10.1177/1403494810395825

28. Kildemoes HW, Sørensen HT, Hallas J. The Danish National Prescription Registry. Scand J Public Health. Jul 2011;39(7 Suppl):38-41. doi:10.1177/1403494810394717

29. Pedersen CB. The Danish Civil Registration System. Scandinavian Journal of Public Health. 2011/07/01 2011;39(7_suppl):22–25. doi:10.1177/1403494810387965

30. Schmidt M, Pedersen L, Sørensen HT. The Danish Civil Registration System as a tool in epidemiology. Eur J Epidemiol. Aug 2014;29(8):541–9. doi:10.1007/s10654-014-9930-3

31. Bybjerg-Grauholm J, Bøcker Pedersen C, Bækvad-Hansen M, et al. 2020;doi:10.1101/2020.11.30.20237768

32. Prive F, Luu K, Blum MGB, McGrath JJ, Vilhjalmsson BJ. Efficient toolkit implementing best practices for principal component analysis of population genetic data. *Bioinformatics (Oxford*, England). Aug 15 2020;36(16):4449–4457. doi:10.1093/bioinformatics/btaa520

33. Manichaikul A, Mychaleckyj JC, Rich SS, Daly K, Sale M, Chen WM. Robust relationship inference in genome-wide association studies. Bioinformatics. Nov 15 2010;26(22):2867–73. doi:10.1093/bioinformatics/btq559

34. Prive F, Arbel J, Vilhjalmsson BJ. LDpred2: better, faster, stronger. *Bioinformatics (Oxford*, England). Dec 16 2020;doi:10.1093/bioinformatics/btaa1029

35. Albinana C, Zhu Z, Schork AJ, et al. Multi-PGS enhances polygenic prediction by combining 937 polygenic scores. Nat Commun. Aug 5 2023;14(1):4702. doi:10.1038/s41467-023-40330-w

36. Larsen JT, Bulik CM, Thornton LM, Koch SV, Petersen L. Prenatal and perinatal factors and risk of eating disorders. Psychol Med. Apr 2021;51(5):870–880. doi:10.1017/s0033291719003945

37. Li Y, Sjolander A, Song H, et al. Associations of parental and perinatal factors with subsequent risk of stress-related disorders: a nationwide cohort study with sibling comparison. Mol Psychiatry. Mar 2022;27(3):1712–1719. doi:10.1038/s41380-021-01406-5

38. Marsál K, Persson PH, Larsen T, Lilja H, Selbing A, Sultan B. Intrauterine growth curves based on ultrasonically estimated foetal weights. Acta paediatrica (Oslo, Norway: 1992). Jul 1996;85(7):843-8. doi:10.1111/j.1651-2227.1996.tb14164.x

39. Persson M, Razaz N, Tedroff K, Joseph KS, Cnattingius S. Five and 10 minute Apgar scores and risks of cerebral palsy and epilepsy: population based cohort study in Sweden. BMJ. Feb 7 2018;360:k207. doi:10.1136/bmj.k207

40. Sedgwick P. Multiple significance tests: the Bonferroni correction. Bmj. 2012;344

41. Sanderson E, Macdonald-Wallis C, Davey Smith G. Negative control exposure studies in the presence of measurement error: implications for attempted effect estimate calibration. International journal of epidemiology. Apr 1 2018;47(2):587–596. doi:10.1093/ije/dyx213

42. Leppert B, Havdahl A, Riglin L, et al. Association of Maternal Neurodevelopmental Risk Alleles With Early-Life Exposures. JAMA Psychiatry. Aug 1 2019;76(8):834–842. doi:10.1001/jamapsychiatry.2019.0774

43. Kearns NT, Carl E, Stein AT, et al. Posttraumatic stress disorder and cigarette smoking: A systematic review. Depress Anxiety. Nov 2018;35(11):1056–1072. doi:10.1002/da.22828

44. Pereira B, Figueiredo B, Miguel Pinto T, Miguez MC. Tobacco consumption from the 1st trimester of pregnancy to 7 months postpartum: Effects of previous tobacco consumption, and depression and anxiety symptoms. Addict Behav. Jan 2022;124:107090. doi:10.1016/j.addbeh.2021.107090

45. Bulik-Sullivan BK, Loh PR, Finucane HK, et al. LD Score regression distinguishes confounding from polygenicity in genome-wide association studies. Nat Genet. Mar 2015;47(3):291–5. doi:10.1038/ng.3211

46. Swanson SA, Tiemeier H, Ikram MA, Hernán MA. Nature as a Trialist?: Deconstructing the Analogy Between Mendelian Randomization and Randomized Trials. Epidemiology. Sep 2017;28(5):653–659. doi:10.1097/ede.0000000000000699

47. Guo W, Guo Y, Song S, et al. Causal effect of the age at first birth with depression: a mendelian randomization study. BMC Medical Genomics. 2024/07/24 2024;17(1):192. doi:10.1186/s12920-024-01966-9

48. Yu Y, Hou L, Wu Y, et al. Causal associations between female reproductive behaviors and psychiatric disorders: a lifecourse Mendelian randomization study. BMC Psychiatry. 2023/11/02 2023;23(1):799. doi:10.1186/s12888-023-05203-y

49. Zhang R, Sjolander A, Ploner A, Lu D, Bulik CM, Bergen SE. Novel disease associations with schizophrenia genetic risk revealed in ∼400,000 UK Biobank participants. Mol Psychiatry. Mar 2022;27(3):1448–1454. doi:10.1038/s41380-021-01387-5

50. Vancampfort D, Stubbs B, Mitchell AJ, et al. Risk of metabolic syndrome and its components in people with schizophrenia and related psychotic disorders, bipolar disorder and major depressive disorder: a systematic review and meta-analysis. World psychiatry: official journal of the World Psychiatric Association (WPA). Oct 2015;14(3):339–47.

51. Nielsen RE, Banner J, Jensen SE. Cardiovascular disease in patients with severe mental illness. Nature reviews Cardiology. Feb 2021;18(2):136–145. doi:10.1038/s41569-020-00463-7

52. Ross LE, Grigoriadis S, Mamisashvili L, et al. Selected pregnancy and delivery outcomes after exposure to antidepressant medication: a systematic review and meta-analysis. JAMA Psychiatry. Apr 2013;70(4):436–43. doi:10.1001/jamapsychiatry.2013.684

53. Huang H, Coleman S, Bridge JA, Yonkers K, Katon W. A meta-analysis of the relationship between antidepressant use in pregnancy and the risk of preterm birth and low birth weight. General hospital psychiatry. Jan-Feb 2014;36(1):13–8. doi:10.1016/j.genhosppsych.2013.08.002

54. Rommel AS, Momen NC, Molenaar NM, et al. Antidepressant use during pregnancy and risk of adverse neonatal outcomes: A comprehensive investigation of previously identified associations. Acta Psychiatr Scand. Jun 2022;145(6):544–556. doi:10.1111/acps.13409

55. Jarde A, Morais M, Kingston D, et al. Neonatal Outcomes in Women With Untreated Antenatal Depression Compared With Women Without Depression: A Systematic Review and Meta-analysis. JAMA Psychiatry. Aug 1 2016;73(8):826–37. doi:10.1001/jamapsychiatry.2016.0934

56. Goodman JH. Women’s attitudes, preferences, and perceived barriers to treatment for perinatal depression. Birth. Mar 2009;36(1):60–9. doi:10.1111/j.1523-536X.2008.00296.x

