## Supplementary for "Association between maternal genome-wide polygenic scores for psychiatric and neurodevelopmental disorders and perinatal risk factors: A Danish population-based study"

**Supplementary Figure 1** The distribution of maternal polygenic scores for psychiatric and neurodevelopmental disorders, comparing those with and without specific disorder diagnoses and antidepressant use in the six months before conception.

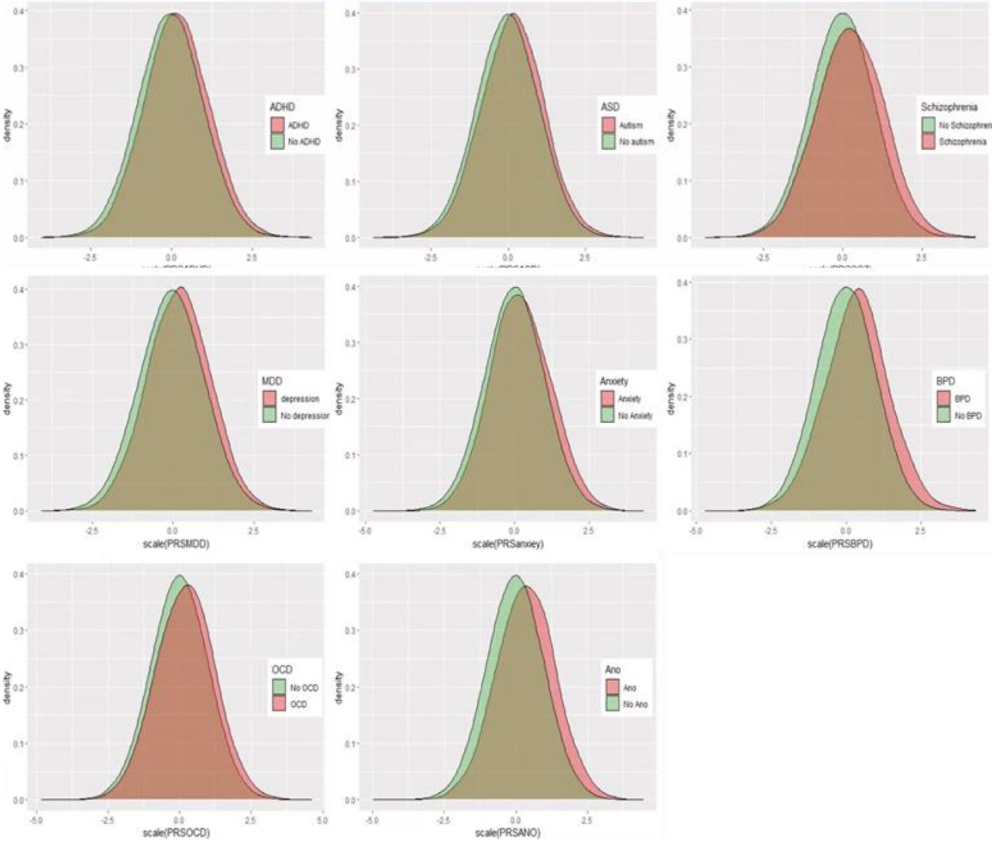

Note, ADHD, Attention-deficit/hyperactivity disorder; ASD, Autism Spectrum Disorder; MDD, Major Depressive Disorder/Depression; BPD, Bipolar disorder; OCD, Obsessive-Compulsive Disorder; ANO, Anorexia Nervosa.

**Supplementary Figure 2** Associations between maternal polygenic scores for psychiatric and neurodevelopmental disorders and perinatal risk factors, among representative population group (n=4115)

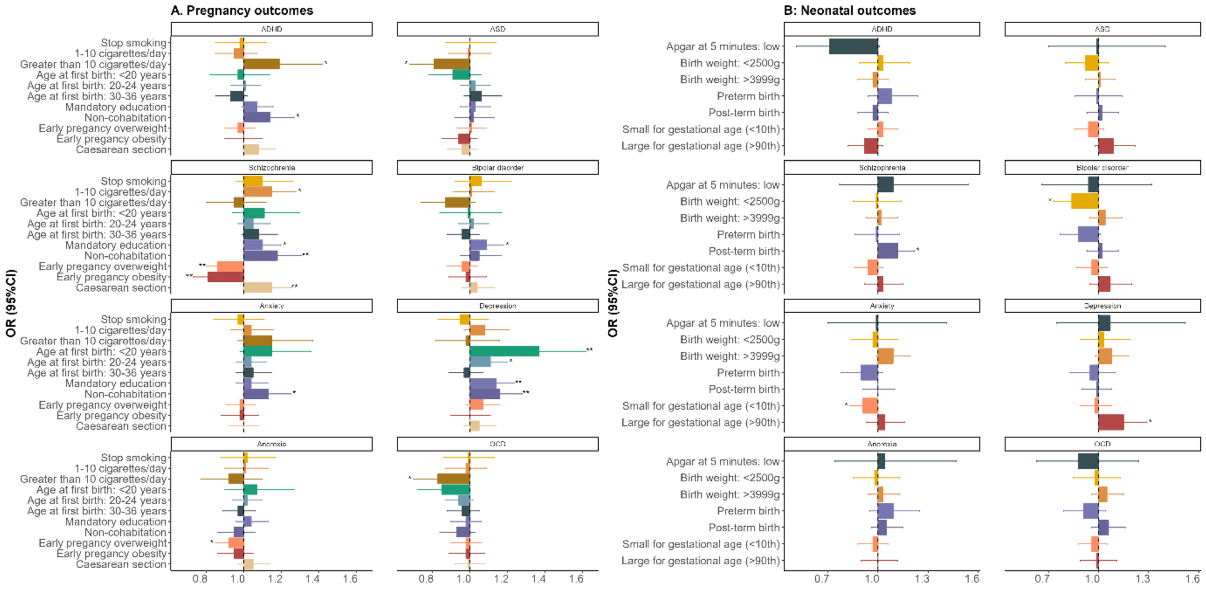

* p < 0.05

** p.adjust < 0.0083 for pregnancy related factors and p.adjust < 0.0125 for neonatal related factors.

**Supplementary table 1** Detailed descriptions of registers used in this study.

| National registry | Year covered | Variables relevant to the current study |
|---|---|---|
| Medical Birth Register | 1997-2015 | Pregnancy and Neonatal factors |
| Psychiatric Central Research Registers | Since 1968 | Mental illness (ICD-10 code, F chapter) |
| Civil Registration System | Since 1968 | Birth date, cohabitation status |
| National Prescription Registry | Since 1995 | Antidepressant (ATC code N06A) |

ICD-10: International Classification of Diseases 10th

**Supplementary table 2** Detailed descriptions of summary statistics used in this study.

| Mental illness | PubMed ID | N | Ncases | Ncontrol |
|---|---|---|---|---|
| ADHD | 30478444 | 15237 | 4225 | 11012 |
| ASD | 30804558 | 10610 | 5305 | 5305 |
| Schizophrenia | Preprint | 132427 | 64689 | 67738 |
| Depression | 30718901 | 775457 | 230118 | 545339 |
| Anxiety | 31427789 | 118397 | 31655 | 86742 |
| Bipolar disoder | Preprint | 385726 | 40488 | 345238 |
| OCD | 28761083 | 9725 | 2688 | 7037 |
| Anorexia nervosa | 31308545 | 45671 | 11940 | 33731 |

Note, ADHD, Attention-deficit/hyperactivity disorder; ASD, Autism Spectrum Disorder; OCD, Obsessive-Compulsive Disorder.

**Supplementary table 3** Associations of maternal PRS for left-handedness (control PRS) and perinatal risk factors (N=7,816).

| Perinatal risk factors | OR (95%CI) | p.value |
|---|---|---|
| **Pregnancy related** | | |
| **Maternal smoking** | | |
| Non-smoker | Ref | |
| Cessation | 1.05 ( 0.96 - 1.15 ) | 0,30 |
| 1-10 cigarettes per day | 0.99 ( 0.93 - 1.06 ) | 0.85 |
| More than 10 cigarettes per day | 0.97 ( 0.88 - 1.06 ) | 0.48 |
| Smoker, unknown cigarettes per day | 0.88 ( 0.85 - 0.91 ) | 0.00 |
| **Maternal BMI at early pregnancy** | | |
| Underweight or normal weight (<25 $kg/m^2$) | Ref | |
| Overweight (25-30 $kg/m^2$) | 0.97 ( 0.91 - 1.03 ) | 0.29 |
| Obesity (⩾30 $kg/m^2$) | 0.96 ( 0.89 - 1.03 ) | 0.23 |
| **Maternal age at first birth, years** | | |
| 25-29 | Ref | |
| <20 | 1.03 ( 0.95 - 1.12 ) | 0.51 |
| 20-24 | 0.98 ( 0.93 - 1.04 ) | 0.47 |
| 30-36 | 1.00 ( 0.93 - 1.09 ) | 0.91 |
| **Maternal cohabitation** | | |
| Cohabitation | Ref | |
| Non-cohabitation | 1.00 ( 0.95 - 1.07 ) | 0.88 |
| **Maternal education** | | |
| More than mandatory education | Ref | |
| Mandatory | 0.99 ( 0.94 - 1.04 ) | 0.68 |
| **Caesarean section** | | |
| No | Ref | |
| Yes | 1.02 ( 0.97 - 1.09 ) | 0.43 |
| **Neonatal related** | | |
| **Birth weight for gestational age** | | |
| Normal for gestation age | Ref | |
| Small for gestational age | 1.01 ( 0.94 - 1.08 ) | 0.76 |
| Large for gestational age | 1.00 ( 0.92 - 1.08 ) | 0.93 |
| **Birth weight** | | |
| 2500-3999g | Ref | |
| <2500g | 1.05 ( 0.95 - 1.15 ) | 0.37 |
| >3999g | 1.02 ( 0.95 - 1.09 ) | 0.55 |
| **Gestational age** | | |
| Term | | |
| Preterm | 1.07 ( 0.97 - 1.18 ) | 0.15 |
| Post.term | 1.01 ( 0.95 - 1.08 ) | 0.72 |
| **Apgar at 5 minutes** | | |
| Normal | Ref | |
| Low | 1.18 ( 0.94 - 1.48 ) | 0.15 |
